# Resting State Cortical Network and Subcortical Hyperconnectivity in Youth With Generalized Anxiety Disorder in the ABCD Study

**DOI:** 10.1101/2024.09.07.24313237

**Authors:** Sam A. Sievertsen, Jinhan Zhu, Angela Fang, Jennifer K. Forsyth

**Author notes:** Correspondence concerning this article should be addressed to Jennifer Forsyth, 4000 15th AVE NE Guthrie Hall 119A, Seattle, WA 98195-0008.

## Abstract

**Introduction:** Generalized anxiety disorder (GAD) frequently emerges during childhood or adolescence, yet, few studies have examined functional connectivity differences in youth GAD. Functional MRI studies of adult GAD have implicated multiple brain regions; however, frequent examination of individual brain seed regions and/or networks has limited a holistic view of GAD-associated differences. The current study therefore used resting-state fMRI data from the Adolescent Brain Cognitive Development study to investigate connectivity in youth with GAD across multiple cortical networks and subcortical regions implicated in adult GAD, considering diagnosis changes across two assessment periods.

**Methods:** Within- and between-network connectivity in 164 GAD youth and 3158 healthy controls for 6 cortical networks and 6 subcortical regions was assessed using linear mixed effect models. Changes in GAD-associated connectivity between baseline and 2-year follow-up were then compared for subjects with: continuous GAD, GAD at baseline and not follow-up (GAD-remitters), GAD at follow-up and not baseline (GAD-converters), and controls. Associations between GAD-associated connectivity metrics and Child Behavior Checklist (CBCL) symptom severity were assessed using mixed effects models.

**Results:** GAD youth showed greater within-ventral attention network (VAN) connectivity, and hyperconnectivity between the amygdala and cingulo-opercular network, and between striatal regions and the cingulo-opercular, default mode, and salience networks (FDR p<0.05). Within-VAN connectivity decreased for GAD-remitters between baseline and follow-up. Connectivity was not associated with symptom severity.

**Discussion:** Results indicate that GAD in childhood and adolescence is associated with altered subcortical to cortical network connectivity affecting multiple networks, and that within-VAN hyperconnectivity, in particular, is associated with clinically-significant GAD symptoms.

## Introduction

Generalized anxiety disorder (GAD) is characterized by diffuse, persistent, and excessive worry across various domains of everyday life (1), and affects 5.7% of individuals throughout their lifetime (2). GAD commonly manifests in late childhood or adolescence (3), a critical period for brain development (4,5). Onset of GAD in childhood or adolescence is associated with a high rate of psychiatric comorbidity, perpetuation of symptoms into adulthood, and increased suicidality risk (6-12). Understanding the neurobiological mechanisms underlying pediatric GAD may improve our ability to treat its core symptoms and reduce the likelihood of psychiatric sequelae.

Functional magnetic resonance imaging (fMRI) studies are key tools for understanding the neurobiology of GAD. In particular, resting-state fMRI (rs-fMRI) studies have shown that distant regions of the brain are co-activated within reproducible intrinsic functional connectivity networks that support distinct aspects of behavior, cognition, and emotion (13-15). Rs-fMRI and task-based fMRI studies of adults with GAD have identified connectivity differences in several cortical networks, including the cingulo-opercular network (CON) (16), involved in tonic arousal and changes in cognitive control (17,18); the default mode network (DMN) (19,20), involved in self-referential processing (21,22); and the frontoparietal network (FPN) (16,23), involved in initiating cognitive control and integrating attention networks (24,25). Abnormalities have also been found in attention-related networks, including the ventral attention network (VAN) (16,26), involved in detection of unexpected but behaviorally relevant stimuli (27,28), the dorsal attention network (DAN) (16), involved in top-down attentional selection (27,28), and the salience network (SN) (29,30), involved in detection of behavioral stimuli and resource direction (31,32). Initial studies of youth with a history of depression or anxiety found altered connectivity involving regions of the VAN and CON (26,33,34); however, connectivity within and between these networks has yet to be systematically assessed in pediatric GAD, specifically.

Additionally, alterations in the structure and/or activity of multiple subcortical regions have been identified in adult GAD. Extant findings have implicated the amygdala (23,35,36), involved in fear and threat detection (37); the hippocampus (38), involved in learning and memory (22,37,39); the thalamus (38,40), considered the brain’s sensory relay center (41-43); and the caudate (44,45), putamen (46,47), and nucleus accumbens (46), which are basal ganglia structures known to form loops with multiple cortical regions to facilitate goal-directed behavior, reward processing, and cognitive functions (48-51). While cortical-amygdala connectivity has been frequently studied in anxiety disorders (52), connectivity with broader subcortical structures has been less studied, despite evidence of structural and intrinsic functional connectivity between multiple subcortical nodes and cortical networks (53,54). Systematic exploration of cortical network-subcortical region connectivity in larger samples of pediatric GAD may offer new insights into connectivity changes that contribute to GAD’s onset and maintenance.

In this study, we therefore leveraged data from the Adolescent Brain and Cognitive Development (ABCD) study, the largest existing study of youth in the United States involving longitudinal clinical, behavioral, and neuroimaging data. By systematically examining functional connectivity within and between cortical rs-fMRI networks and subcortical regions implicated in previous fMRI studies of adult GAD, we aim to comprehensively map connectivity alterations associated with a GAD diagnosis during childhood or early adolescence. Furthermore, leveraging the longitudinal nature of ABCD study data, we aim to identify whether connectivity changes in cortical networks or between cortical networks and subcortical regions are associated with the emergence or remission of a GAD diagnosis over time. Mapping these alterations in youth with GAD may provide insights into key networks to target for earlier intervention.

## Methods and Materials

### Participants

Data for 11,878 subjects recruited at ages 8-11 from 21 research sites across the United States available from data release version 4.0 of the ABCD study was used for the current analyses. Rs-fMRI data was available at baseline, when youth were 8-11 years of age and 2-year follow-up when youth were 10-13 years of age. To maximize sample size of youth with GAD for the present analyses, rs-fMRI data was included for youth who met criteria for current GAD at the baseline or 2-year follow-up time-point (n=164). For GAD subjects who met disorder criteria at both time-points, we randomly selected which time-point to use. Healthy control (HC) comparison subjects were identified as those who did not meet lifetime criteria for any assessed mental health diagnoses at either time-point. To account for potential confounds related to assessment time-point and site, rs-fMRI data used for HC subjects was selected across time-points to be proportionally matched to the assessment time-point used for GAD subjects, within each site, leaving a final sample of 3158 HC and 164 GAD subjects for primary analyses (see Table 1). Data was accessed from the National Institute of Mental Health Data Archive (dataset: https://doi.org/10.15154/1523041).

**Table 1:**
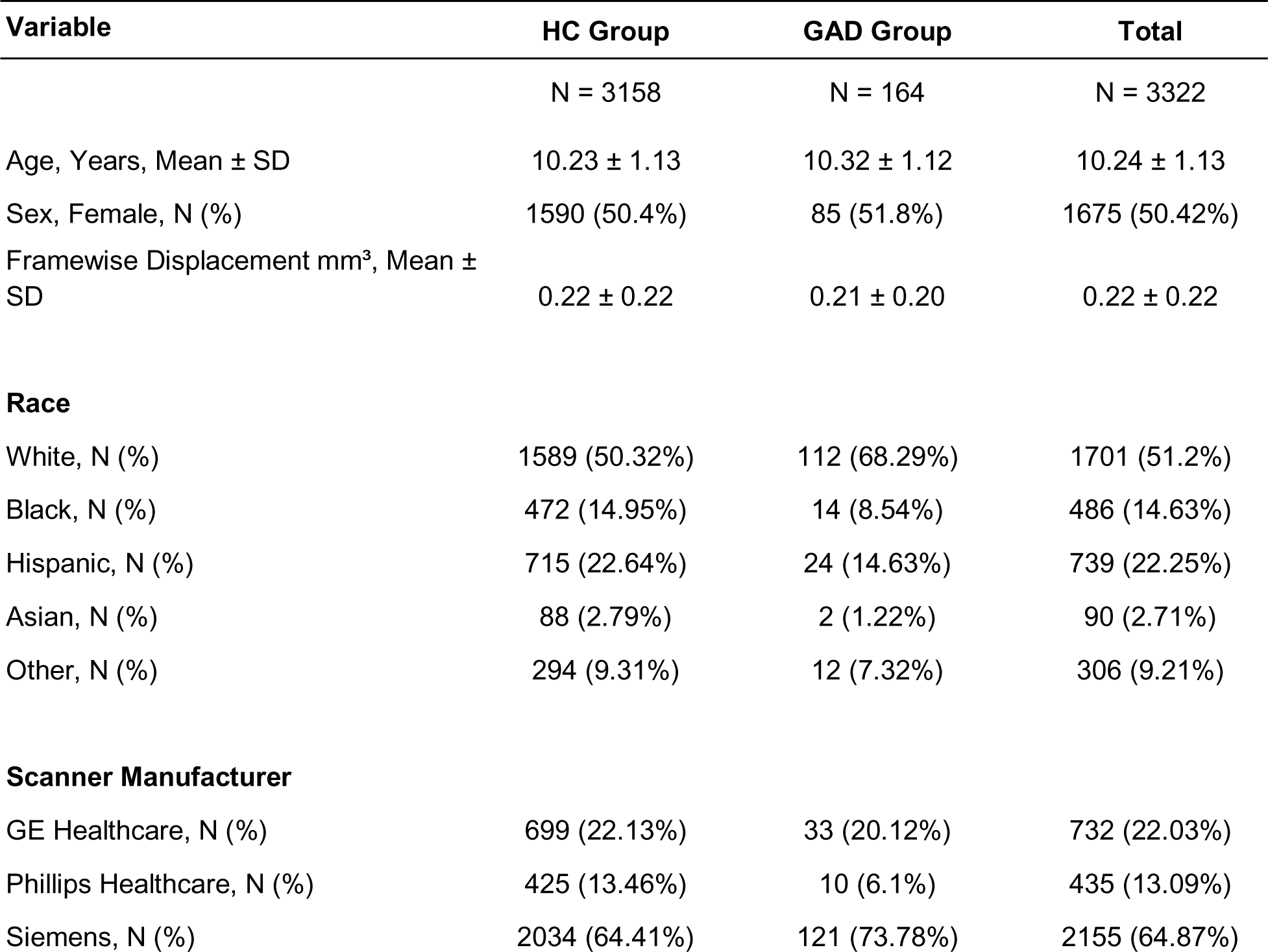
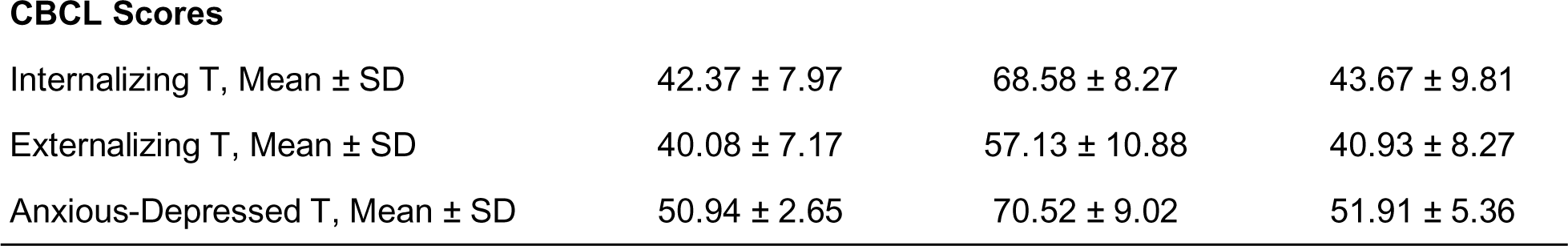
Characteristics of the Sample.

| Variable | HC Group | GAD Group | Total |
| --- | --- | --- | --- |
|  | N = 3158 | N = 164 | N = 3322 |
| Age, Years, Mean ± SD | 10.23 ± 1.13 | 10.32 ± 1.12 | 10.24 ± 1.13 |
| Sex, Female, N (%) | 1590 (50.4%) | 85 (51.8%) | 1675 (50.42%) |
| Framewise Displacement mm<sup>3</sup>, Mean ± SD | 0.22 ± 0.22 | 0.21 ± 0.20 | 0.22 ± 0.22 |
| **Race** |  |  |  |
| White, N (%) | 1589 (50.32%) | 112 (68.29%) | 1701 (51.2%) |
| Black, N (%) | 472 (14.95%) | 14 (8.54%) | 486 (14.63%) |
| Hispanic, N (%) | 715 (22.64%) | 24 (14.63%) | 739 (22.25%) |
| Asian, N (%) | 88 (2.79%) | 2 (1.22%) | 90 (2.71%) |
| Other, N (%) | 294 (9.31%) | 12 (7.32%) | 306 (9.21%) |
| **Scanner Manufacturer** |  |  |  |
| GE Healthcare, N (%) | 699 (22.13%) | 33 (20.12%) | 732 (22.03%) |
| Phillips Healthcare, N (%) | 425 (13.46%) | 10 (6.1%) | 435 (13.09%) |
| Siemens, N (%) | 2034 (64.41%) | 121 (73.78%) | 2155 (64.87%) |
| CBCL Scores |  |  |  |
| Internalizing T, Mean ± SD | 42.37 ± 7.97 | 68.58 ± 8.27 | 43.67 ± 9.81 |
| Externalizing T, Mean ± SD | 40.08 ± 7.17 | 57.13 ± 10.88 | 40.93 ± 8.27 |
| Anxious-Depressed T, Mean ± SD | 50.94 ± 2.65 | 70.52 ± 9.02 | 51.91 ± 5.36 |

### Measures

Current and lifetime history of psychiatric disorders were determined using the parent report of the computerized Kiddie Schedule for Affective Disorders and Schizophrenia for DSM-5 (KSADS-COMP) structured clinical interview (55-57).

Child Behavior Checklist (CBCL) scores were used to capture dimensional symptom severity/frequency within the last 6 months (58). We prioritized the parent report as it was available and complete for the greatest number of subjects at baseline and 2-year follow-up. For this study, T-scores for the anxious-depressed, internalizing, and externalizing dimensions were used. CBCL scores were available for 157 GAD subjects and 2989 HC subjects.

### Imaging procedure

The neuroimaging protocol harmonized across scanners at each of the 21 sites in the ABCD study has been detailed elsewhere (59). Briefly, youth were scanned in a 3-T scanner (Siemens, General Electric, or Philips model) with a 32-channel head coil. Participants underwent four 5-minute blood oxygen level dependent (BOLD) rsfMRI scans per assessment, with their eyes open and fixated on a static crosshair. BOLD rsfMRI images were acquired in the axial plane using an echo planar imaging sequence. To meet quality control (QC) criteria for this study, both the rsfMRI image series and T1-weighted imaging series needed to pass raw QC, and the number of usable rs-fMRI frames after censoring needed to be >375. For additional information on how framewise and whole scan motion, distortion, and other signals of non-interest were accounted for, see (60). Subcortical regions were parcellated using Freesurfer’s automatic subcortical segmentation atlas (61) and functionally-defined cortical network ROI were parcellated according to the Gordon atlas (62). Cortical network connectivity metrics were calculated as the pairwise mean pearson correlations within and between networks. These correlation coefficients were Fisher-transformed into z-scores, and averaged to measure network correlation strength. Metrics for mean pairwise correlation between cortical networks of interest and between cortical networks and subcortical regions were also utilized. Primary analyses focused on within- and between-network connectivity for 6 cortical networks implicated in adult GAD: the DMN, SN, VAN, DAN, CON, and FPN cortical networks (22 pairwise comparisons). Additionally, connectivity between each cortical network and 6 subcortical regions (left and right hemisphere) implicated in adult GAD were investigated, specifically, the thalamus, caudate, nucleus accumbens, putamen, hippocampus, and amygdala (72 pairwise comparisons), yielding 94 total pairwise functional connectivity metrics.

## Statistical Analysis

### Overall HC Versus GAD Group Differences in Connectivity

Analyses were conducted in R version 4.2.2 (63). Connectivity for the 94 metrics of interest were modeled across GAD and HC subjects using linear mixed-effect models from the *lmerTest* package (64), with group and covariates for biological sex, assessment time-point, and mean framewise displacement in mm³ (FD) specified as fixed effects. Scanner and family ID were modeled as random effect covariates in accordance with analysis recommendations for the ABCD study (65-68); GAD vs. HC group differences were tested using type II analysis of covariance (ANCOVA; Wald F tests with Kenward-Roger degrees of freedom (69)) from the *car* package (70). To reduce type I error, p-values for group effects were adjusted for multiple testing across the 94 connectivity metrics of interest using the Benjamini & Hochberg false discovery rate (FDR) (71) correction. Rs-fMRI metrics associated with GAD after multiple-testing correction were retained for further downstream analyses.

### Change in GAD-Associated Connectivity Metrics Across Time

To further understand the relationship between GAD diagnostic status and connectivity differences, we examined changes in GAD-associated connectivity metrics as subjects developed or remitted from a GAD diagnosis between baseline and 2-year follow-up. Specifically, subjects with both baseline and 2-year follow-up rs-fMRI data available were grouped into 4 subgroups: 1) subjects who met criteria for GAD at both time-points (i.e., a continuous GAD group, *n=*12); 2) subjects who met criteria for current GAD diagnosis at baseline but not at follow-up (i.e., a GAD remitter group; *n*=52); 3) subjects who met current GAD criteria at follow-up but not at baseline (i.e., a GAD converter group; *n*=55), and 4) continuously healthy controls (*n*=1955). Change in connectivity from baseline to 2-year follow-up was modeled using linear mixed-effects models, with diagnostic subgroup, assessment time-point, mean FD, and biological sex modeled as fixed effects, and subject, scanner, and family ID modeled as random effects. An interaction term was specified between diagnostic subgroup and assessment time-point. Potential interactions between subgroups and connectivity over time were then tested for significance using a type III ANCOVA from the *car* package^44^. Resulting p-values from the subgroup by time interaction across GAD-associated connectivity metrics investigated were FDR corrected. Subsequently, for models showing nominally significant interactions between subgroup and time, estimated marginal means were computed and used to examine linear trends across time within groups and differences between groups in connectivity slopes from baseline to the 2-year follow-up time-point.

### GAD-Associated Connectivity Metrics Versus Dimensional Symptoms

To assess the relationship between symptom severity and observed connectivity patterns, we examined relationships between CBCL T-scores for internalizing, externalizing, and anxious-depressed symptoms and GAD-associated connectivity metrics using linear mixed-effects models with biological sex, assessment time-point, and mean FD in mm³ as fixed effect covariates, and scanner and family ID as random effect covariates. These relationships were first assessed among youth with active GAD diagnoses only and were then expanded to include GAD and HC subjects, specifying diagnostic group as an interaction term with dimensional symptoms. A type III ANCOVA was used to test the omnibus interaction effect between diagnostic group and CBCL score in their association with connectivity.

Parallel sensitivity analyses were conducted with the abbreviated youth-self report version of the CBCL, the youth brief problem monitor (BPM) (72) available for a subset of subjects at 2-year follow-up (GAD *n=*46, HC *n=*928).

## Results

### Group Differences in Cortical Network and Subcortical Region Connectivity

There were significant group differences in functional connectivity for 5 metrics. Specifically, GAD youth displayed significant within-VAN hyperconnectivity (F(1,3245.88)=9.3, p=0.002, FDR p=0.044, Figure 1A), as well as hyperconnectivity between the CON and left amygdala (F(1,3238)=11.85, p<.001, FDR p=0.028), CON and left caudate (F(1,3241.19)=9.3, p=0.002, FDR p=0.044), DMN and left putamen (F(1,3253.87)=9.85, p=0.002, FDR p=0.044), and SN and left putamen (F(1,3260.12)=14.38, p<.001, FDR p=0.014, Figure 1B-E) compared to HC. There were no significant differences reflecting reduced connectivity in the GAD group.

**Figure 1:**
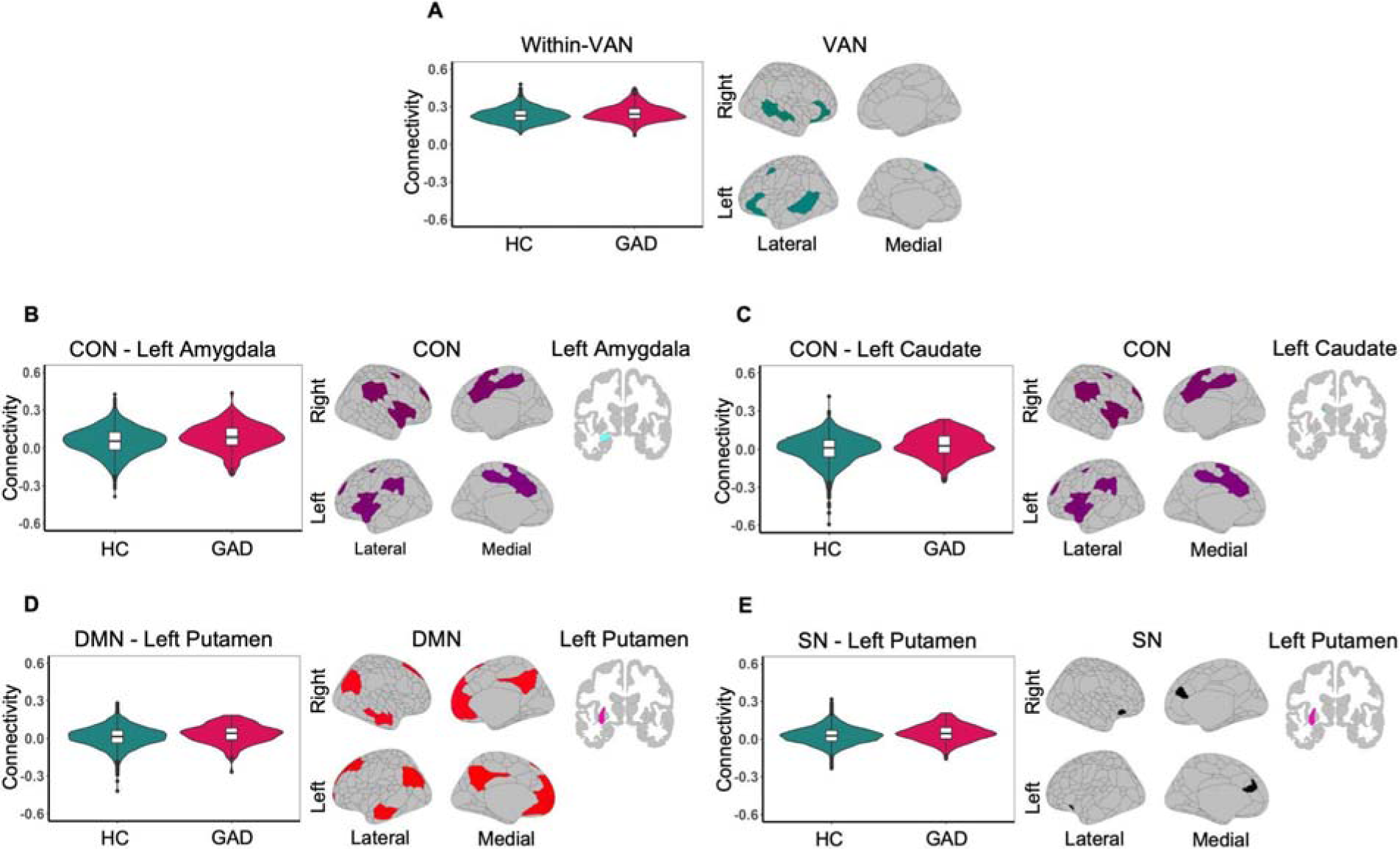
Group differences in within-cortical network connectivity and cortical network - subcortical region connectivity metrics for GAD youth as compared to HC (all FDR p<0.05). (**A**) GAD group within-ventral attention network (VAN) hyperconnectivity. (**B**) GAD group cingulo opercular network (CON) - left amygdala hyperconnectivity. (**C**) GAD group CON - left caudate hyperconnectivity. (**D**) GAD group default mode network (DMN) - left putamen hyperconnectivity. (**E**) GAD group salience network (SN) - left putamen hyperconnectivity.

Nominal GAD group hyperconnectivity was also observed between the CON and VAN (F(1,3251.26)=5.65, p=0.018, FDR p=0.183), partially replicating a recent finding linking CON-VAN hyperconnectivity to heightened anxiety symptoms (34), between the CON and SN (F(1,3255.84)=4.74, p=0.03, FDR p=0.214), and between the CON, DMN, DAN, SN and multiple subcortical regions (all p’s <0.05); however, these associations did not survive multiple-testing correction. Results for all 94 connectivity models are provided in Table S1.

### Changes in GAD Diagnosis Status and Connectivity Over Time

The majority of subjects with a current GAD diagnosis at either baseline or year 2 follow-up experienced a change in GAD status between assessment visits. Among the 5 GAD-associated connectivity metrics described above, between baseline and 2-year follow-up there were no significant interactions between GAD subgroup and time-point for the CON - left amygdala, CON - left caudate, DMN - left putamen, and SN - left putamen connectivity metrics. However, there was a nominally significant interaction between GAD subgroup and assessment time-poin for within-VAN connectivity (F(3,2070)=2.87, p=0.035, FDR p=0.176, Table S2). Follow-up analyses of within group changes from baseline to 2-year follow-up revealed a significant decrease in within-VAN connectivity for both controls (b=-0.006, 95% CI [0.004, 0.009], p<.001) and GAD remitters (b=-0.026, 95% CI [0.01, 0.042], p=0.001). Within-VAN connectivity non-significantly increased between assessments for the GAD converters and non-significantly decreased for continuous GAD subjects (p>0.05; Figure 2). Additionally, pairwise comparison of linear contrasts from baseline to 2-year follow-up between groups suggested that GAD remitters showed a greater decrease in within-VAN connectivity compared to controls (p=0.02, FDR p=0.055) and GAD converters (p=0.013, FDR p=0.055), although these pairwise comparisons did not survive multiple-testing correction.

**Figure 2:**
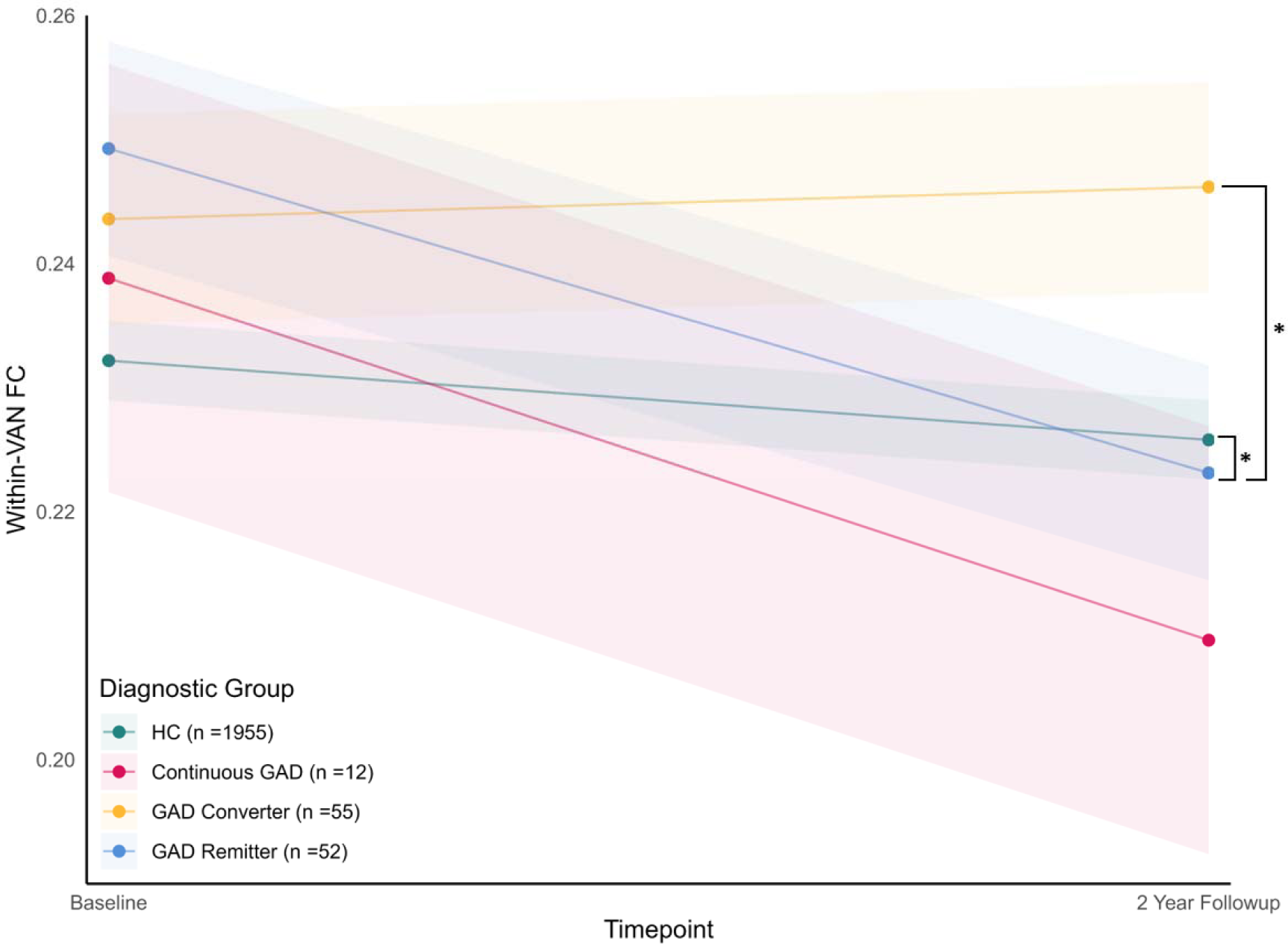
Change in within-ventral attention network (VAN) functional connectivity (FC) from baseline to 2-year follow-up for GAD subgroups and healthy controls (HC). *Pairwise linear slopes contrast between groups p<0.05.

### Associations Between Dimensional Symptoms and Connectivity

Among subjects with current GAD, functional connectivity was not significantly associated with severity of anxious-depressed, internalizing, or externalizing symptoms from the CBCL. (Table S3; Figure S1). Extending analyses to the full sample, controlling for diagnostic group, there were no significant associations between anxious-depressed or internalizing symptoms and GAD-associated connectivity metrics, nor significant interactions between CBCL scores and diagnostic group on GAD-associated connectivity metrics (Table S4; Figure S2).

Interestingly, nominal associations between anxiety-relevant CBCL scores and GAD-associated connectivity metrics were observed in unexpected directions. Specifically, across the whole sample, higher anxious depressed symptoms were nominally associated with lower within-VAN connectivity (b=-0.001, 95% CI [-0.002, 0], p=0.049, FDR p=0.369). Higher externalizing symptoms were also significantly associated with lower CON - left amygdala connectivity (b=-0.001, 95% CI [-0.001, 0], p=0.002, FDR p=0.030). Exploratory analyses in the HC group alone, which was 20 times the size of the GAD group, suggested that these associations were driven by effects in HCs (see Table S5; Figure S3).

Using the youth BPM showed a similar pattern of results, with no significant associations between anxious-depressed, internalizing or externalizing symptoms and GAD-associated connectivity metrics among subjects with current GAD (Table S6), and nominally significant associations between externalizing symptoms and CON - left amygdala connectivity (b=-0.002, 95% CI [-0.004, 0], p=0.044, FDR p=0.129), CON - left caudate connectivity (b=-0.003, 95% CI [-0.005, -0.001], p=0.006, FDR p=0.055), and DMN - left putamen (b=-0.002, 95% CI [-0.003, 0], p=0.024, FDR p=0.121) in the full sample (Table S7), that appeared to be driven by relationships within the large HC group (Table S8).

## Discussion

In the current study, we observed significant differences in functional connectivity in GAD compared to HC youth. Specifically, GAD youth exhibited hyperconnectivity within the VAN, as well as hyperconnectivity between the CON and left amygdala, CON and left caudate, DMN and left putamen, and SN and left putamen. Notably, within-VAN connectivity decreased from baseline to follow-up in youth who experienced a remission in GAD diagnosis from baseline and follow-up. Furthermore, GAD remitters showed a greater reduction in within-VAN connectivity across time-points compared to GAD converters and HC. However, among youth with a current GAD diagnosis, severity of anxious-depressed symptoms, as measured by the CBCL, was not associated with connectivity in the VAN or any other GAD-associated connectivity metric. Taken together, this suggests a key role for within-VAN hyperconnectivity in the experience of GAD-specific symptoms, independent from severity of broader anxious-depressed symptoms.

A core tenet of GAD is excessive or uncontrollable worry and anxiety across a range of experiences and events (73,74). Identifying the neurobiological basis of this experience may help clarify targets for intervention and improve our ability to prevent psychiatric sequelae for youth with GAD. Importantly, the VAN is known to be involved in involuntary attentional capture (75) and orientation of attention towards unexpected, behaviorally-relevant stimuli (27,28). To our knowledge, no prior studies have examined VAN connectivity in youth with GAD in particular. However, existing research on youth with a history of mood and anxiety symptoms has implicated VAN connectivity alterations (26). Specifically, one previous study of youth with a history of depression or anxiety and healthy youth, found that connectivity between regions of the VAN was positively correlated with dimensional attention bias towards threat, with decreased connectivity between regions of the VAN in youth with a history of depression or anxiety(26). A more recent study on attentional capture in youth with anxiety found that perturbations of involuntary attentional capture while viewing non-threatening stimuli were associated with higher clinician rated anxiety and activity in regions of the VAN (33). Although the current study differed from these prior studies in its focus on connectivity during rs-fMRI versus task-based fMRI, as well as its focus on youth with current GAD versus broader anxiety and depression, our finding of within-VAN hyperconnectivity in youth GAD builds on these prior findings and suggests a core role for the VAN in pediatric anxiety.

Correspondingly, within-VAN hyperconnectivity disappeared in youth who remitted from GAD between baseline and 2-year follow-up. Although the increase in within-VAN hyperconnectivity over time was not significant in youth who developed GAD by 2-year follow-up, this may reflect the presence of subthreshold symptoms at baseline for GAD converters and/or interactions with normative developmental changes in within-VAN connectivity. Indeed, HC youth also showed a significant decrease in within-VAN connectivity from baseline to 2-year follow-up, though significantly less than that observed in GAD remitters. This reduction in within-VAN connectivity in HC from late childhood to early adolescence opposes patterns of increased within-network connectivity with maturation for some intrinsic connectivity networks (76), but is consistent with prior reports for the VAN (27,77). Overall, the observed diagnosis-dependent shifts in within-VAN hyperconnectivity as youth develop or remit from GAD suggests that within-VAN hyperconnectivity may be a key neurobiological basis for hypervigilance to both threatening and non-threatening stimuli (33), and that within-VAN hyperconnectivity could serve as an indicator of clinically significant GAD symptoms.

While rs-fMRI studies of intrinsic functional connectivity networks often use cortical-focused parcellations, anatomical tracing studies in primates and rodents have established that regions of the basal ganglia connect with the thalamus and multiple cortical regions in cortico-striato-thalamo-cortical loops to regulate reward, cognitive, and sensorimotor information processing, and facilitate goal-directed behavior (78-80). Interestingly, in this study, youth with GAD showed hyperconnectivity between the CON and left caudate, the SN and left putamen, and the DMN and the putamen. The caudate and putamen are basal ganglia structures known to connect to regions of the anterior cingulate (81), which is part of both the CON and SN, and the superior frontal gyrus (82), which is part of the CON. Conversely, intrinsic connectivity between the DMN and putamen was previously identified as anti-correlated in an adult sample from the Human Connectome Project (83). To the best of our knowledge, these observations represent novel hyperconnectivity relationships documented in pediatric GAD, and are in line with growing calls to investigate cortico-striatal connectivity in adolescent anxiety disorders (84). Given the roles of the SN, CON, and DMN in reward processing, tonic arousal, and self-referential processing, respectively, our findings suggest altered striatal modulation of these processes in pediatric GAD. Further investigation is warranted to replicate and clarify the implications of these findings.

Beyond evidence of corticostriatal hyperconnectivity in youth with GAD, we observed hyperconnectivity between the CON and amygdala in pediatric GAD. Notably, anti-correlated connectivity patterns have been previously found between the CON and the centromedial, basolateral, and superficial amygdala in healthy individuals (85). Additionally, a previous study of adults with GAD found decreased bilateral amygdala connectivity with regions of the CON. This opposite connectivity pattern in adult GAD compared to the current study may reflect differences depending on age and/or the investigation of individual CON regions versus the entire CON (23). Amygdala-CON hyperconnectivity observed in the current study appears to reflect reduced amygdala-CON anti-correlation in youth with GAD. Although speculative, this is consistent with the amplified response to perceived threats and heightened difficulty in down-regulating threat responses commonly observed in and consistent with cognitive and behavioral models of GAD (86-88).

While hyperconnectivity within the VAN and between multiple cortical networks and subcortical nodes was associated with current GAD diagnosis, these connectivity metrics were not associated with anxious-depressed symptom severity on the CBCL in youth with GAD. Interestingly, a previous study of children with a history of depression and/or anxiety or no psychiatric history similarly found that within-VAN connectivity was not associated with depressive, internalizing, or externalizing symptom severity (26). This lack of association between connectivity metrics and symptom severity could reflect lower specificity of the CBCL anxious-depressed subscale to GAD-specific symptoms or differences in time-scale between symptom reporting period for the CBCL (i.e., 6-months) versus the KSADS for diagnoses (i.e., 2-weeks). Alternatively, functional connectivity during rs-fMRI may be less sensitive to symptom-relevant connectivity differences than task-based fMRI involving anxiety-eliciting tasks. Nevertheless, our findings suggest a threshold effect, wherein within-VAN hyperconnectivity may at least partially underlie the expression of clinically significant GAD symptoms. Further investigation is needed to clarify the relationships between dimensional anxiety and connectivity metrics among youth with GAD.

Despite the insights gained from our study, several limitations should be considered. First, rs-fMRI data may not fully capture neurobiological signatures of all symptoms associated with GAD. For example, a previous task-based fMRI study of youth found that stimulus-driven attentional control was associated with increased connectivity between the CON and VAN (34). Hyperconnectivity between the CON and VAN was only nominally associated with GAD in the current study. Examination of connectivity between these networks using paradigms that more directly index cognitive and behavioral processes associated with GAD symptoms in youth may elicit stronger group differences. In addition, although this is one of the largest reported neuroimaging studies of youth GAD, our clinical sample sizes, and particularly that of the continuous GAD subgroups, were limited. The current study also only included two time-points of data and assessments occurred across a developmental period associated with substantial network maturation. To our knowledge, no other study exists that has identified connectivity differences associated with the development or remittance of a GAD diagnosis in youth.

However, future analyses in larger clinical cohorts and across longer follow-up periods will be useful to further characterize connectivity alterations associated with GAD onset and remission throughout development. Additionally, assessing connectivity in the context of emerging models of brain age may clarify alterations in pediatric GAD and reveal more specific developmental implications (89,90). Finally, connectivity analyses were limited to one parcellation of large-scale, cortical networks and gross subcortical structures. While investigating commonly-used macroscale cortical and subcortical parcellations facilitated the investigation of cortical networks and subcortical regions broadly implicated in adult anxiety disorders and was useful for generating novel insights into cortical-subcortical connectivity in youth GAD, future studies using alternate parcellations and/or more fine-grained cortical and subcortical region analyses may yield further insights into the neurobiological underpinnings of pediatric GAD.

In summary, the current study identified distinct patterns of functional connectivity associated with GAD during late childhood or early adolescence, including within-VAN hyperconnectivity, and hyperconnectivity between the caudate and putamen and multiple cortical networks, and the CON and amygdala. Our finding that within-VAN hyperconnectivity tracked with changes in GAD status across time suggests that within-VAN hyperconnectivity may play an important role in the manifestation of clinically significant GAD symptoms, and is in line with prior hypotheses of a role for the VAN in anxiety disorders. However, connectivity between broader cortical networks and subcortical regions, including the CON, striatum, and amygdala, also appear to contribute to cognitive and emotional alterations in GAD. Overall, results implicate VAN hyperconnectivity as a key neurobiological target for intervention in pediatric GAD, and also underscore the role of a distributed set of cortical networks and subcortical nodes in pediatric GAD.

## Data Availability

Data used in this study was obtained from the Adolescent Brain Cognitive Development (ABCD) Study (https://abcdstudy.org), held in the NIMH Data Archive (NDA) collection #1299. Specifically, the ABCD data used in this report was retrieved from data release 4.0 (https://doi.org/10.15154/1523041).

## Acknowledgements

Data used in this study was obtained from the Adolescent Brain Cognitive Development (ABCD) Study (https://abcdstudy.org), held in the NIMH Data Archive (NDA). Specifically, the ABCD data used in this report was retrieved from data release 4.0 (https://doi.org/10.15154/1523041).

We would like to thank the Center for Social Science Computation and Research at the University of Washington for valuable computational and statistical programming support.

