## Supplementary Figures for "Resting State Cortical Network and Subcortical Hyperconnectivity in Youth With Generalized Anxiety Disorder in the ABCD Study"

**Supplemental Figures**

**Figure S1:** Within GAD group associations between CBCL anxious depressed, internalizing, and externalizing T-scores and GAD-associated functional connectivity metrics.
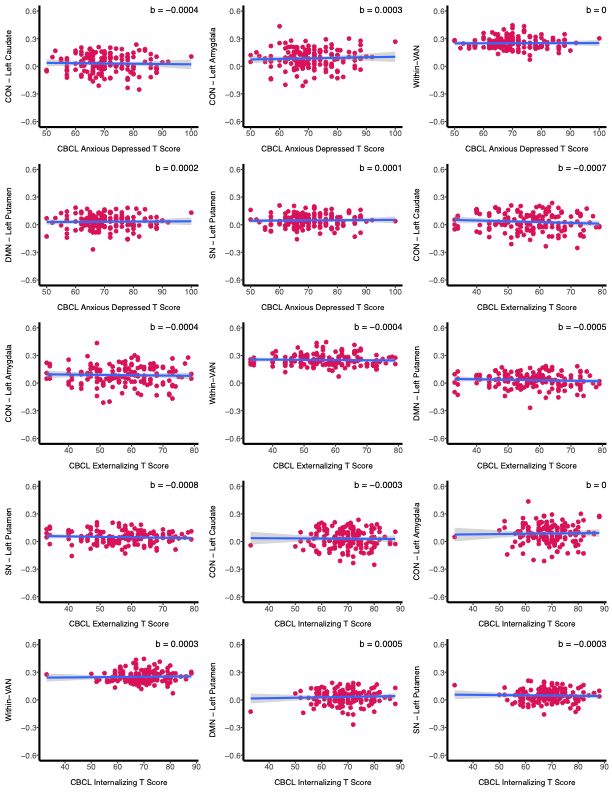

**Figure S2:** Associations between CBCL anxious depressed, internalizing, and externalizing T-scores and GAD-associated functional connectivity metrics across GAD and HC subjects.

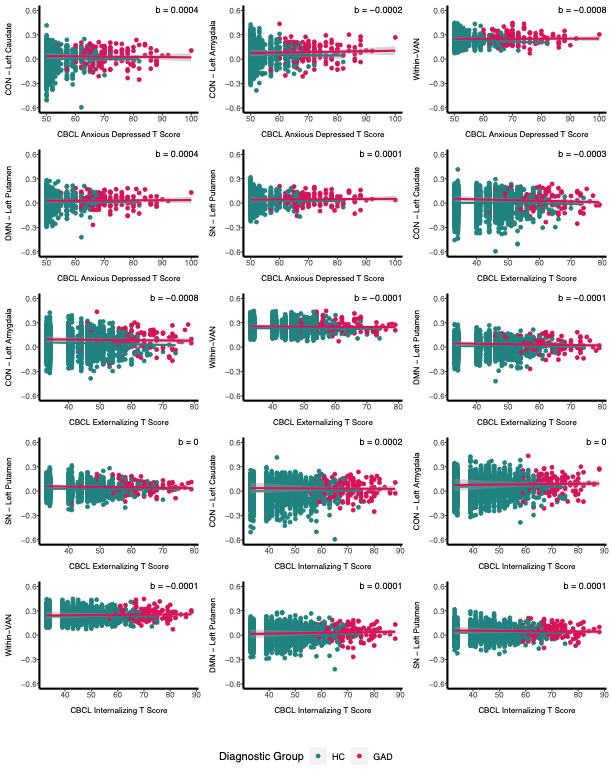

**Figure S3:** Within control group associations between CBCL anxious depressed, internalizing, and externalizing T-scores and GAD-associated functional connectivity metrics.
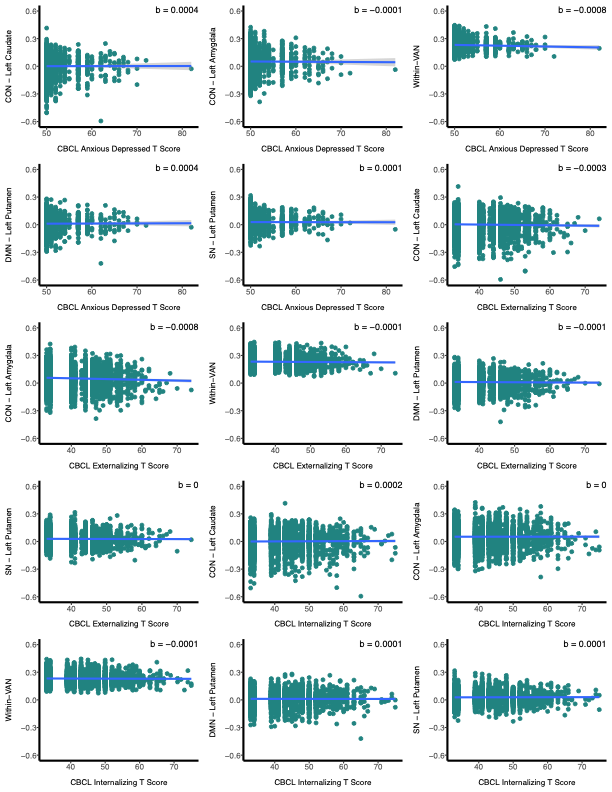
